# Spike ripples localize the epileptogenic zone better than other leading biomarkers: a multicenter intracranial study

**DOI:** 10.1101/2023.04.25.23289111

**Authors:** Wen Shi, Dana Shaw, Katherine G. Walsh, Xue Han, Uri T. Eden, Robert M. Richardson, Stephen V. Gliske, Julia Jacobs, Benjamin H. Brinkmann, Gregory A. Worrell, William C. Stacey, Mark A. Kramer, Catherine J. Chu

**Affiliations:** Department of Neurology, Massachusetts General Hospital, Boston, Massachusetts, USA; Harvard Medical School, Boston, Massachusetts, USA; Graduate Program in Neuroscience, Boston University, Boston, Massachusetts, USA; Center for Systems Neuroscience, Boston University, Boston, Massachusetts, USA; Department of Biomedical Engineering, Boston University, Boston, Massachusetts, USA; Department of Mathematics and Statistics, Boston University, Boston, Massachusetts, USA; Department of Neurosurgery, Massachusetts General Hospital, Boston, Massachusetts, USA; Department of Neurosurgery, University of Nebraska Medical Center, Omaha, Nebraska, USA; Department of Neuropediatrics and Muscle Disorders, Medical Center-University of Freiburg, Freiburg, Germany; Department of Paediatrics, Cumming School of Medicine, University of Calgary, Calgary, AB, Canada; Department of Neuroscience, Cumming School of Medicine, University of Calgary, Calgary, AB, Canada; Hotchkiss Brain Institute and Alberta Children’s Hospital Research Institute, University of Calgary, Calgary, AB,Canada; Bioelectronics Neurophysiology and Engineering Lab, Mayo Clinic, Rochester, Minnesota, USA; Department of Neurology, University of Michigan, Ann Arbor, MI, USA

**Author notes:** Address correspondence to Drs. Shi and Chu, 100 Cambridge Street, Boston, MA 02114.,.

## Abstract

**Objective:** We evaluated whether the combination of epileptiform spikes and ripples (spike ripples) outperformed other leading biomarkers in identifying the epileptogenic zone across subjects in a multicenter international study.

**Methods:** We validated and applied an automated spike ripple detector on intracranial EEG recordings in subjects from 4 centers who subsequently underwent surgical resection with known 1-year seizure outcomes. We evaluated the spike ripple rate in subjects cured after resection (ILAE 1 outcome) and those with persistent seizures (ILAE 2-5) across sites and recording types. We also evaluated spike, wideband HFO (80-500 Hz), fast ripple (250-500 Hz), and ripple (80-250 Hz) rates using validated automated detectors. The proportion of resected events was computed and compared across subject outcomes and biomarkers.

**Results:** 109 subjects were included. The majority of spike ripples were removed in subjects with ILAE 1 outcome (p = 1e-6), and this was qualitatively observed across the four sites (p = 0.032, p = 0.092, p = 0.0005, p = 0.003) and the two electrode types (p = 0.01, p = 7e-6). A higher proportion of spike ripples were removed in subjects with ILAE 1 outcomes compared to ILAE 2-5 outcomes (p = 0.02). Among ILAE 1 subjects, the proportion of spike ripples removed was higher than the proportion of spikes (p = 0.0004), wideband HFOs (p = 0.0004), fast ripples (p = 0.008), and ripples (p = 0.008) removed. At the individual level, more subjects with ILAE 1 outcome had the majority of spike ripples removed (40/48, 83%) than spikes (69%, p = 0.04), wideband HFOs (63%, p = 0.009), fast ripples (36%, p = 2e-5), or ripples (45%, p = 0.0007) removed.

**Interpretation:** When surgical resection was successful, the majority of spike ripples were removed. Automatically detected spike ripples have improved specificity for epileptogenic tissue compared to spikes, wideband HFOs, fast ripples, and ripples.

## Introduction

Epilepsy is the most common neurological disorder, affecting 50 million individuals globally and accounting for 1% of the global disease burden ^1^. One-third of people with epilepsy have persistent seizures despite pharmacological interventions. For subjects with drug-refractory epilepsy, neurosurgical interventions, including resection and neuromodulation, are the most effective treatments. The success of both surgical interventions relies on accurately identifying the brain tissue responsible for generating seizures, known as the epileptogenic zone (EZ).

Interictal spikes and high-frequency oscillations (HFOs) are two widely used biomarkers to identify the EZ ^1, 2^. These biomarkers have been extensively studied but have shown poor specificity in clinical practice. Spikes – high amplitude, sharp, voltage fluctuations – are specific for pathology, but not for the EZ ^3, 4^. Ripples – brief, low-amplitude bursts between 80-250 Hz – are spatially focal ^5^, but are not specific for pathology since similar physiological events are present during normal cognitive processes ^6-8^. Higher frequency (>250 Hz) fast ripples correlate with the epileptogenic zone in subjects with epilepsy but are less frequently observed ^9-11^. Although epileptiform spikes and ripples are understood to represent separate neurophysiological events, nearly half of ripples co-occur with spikes ^12-16^. By combining the pathological specificity of spikes and spatial specificity of ripples, the co-occurrence of these events -- spike ripples -- may yield a more specific biomarker for the EZ compared to either biomarker alone ^7, 17-24^.

To evaluate whether spike ripples provide a reliable and improved biomarker for the EZ compared to other leading biomarkers, we tested three main hypotheses. First, that the majority of automatically detected spike ripples would be removed in subjects who were seizure free after epilepsy surgery. Second, that the subjects who were seizure free after resection would have a higher proportion of spike ripples removed compared to those who were not seizure free. Third, that the subjects who were seizure free after resection would have a higher proportion of spike ripples removed compared to the proportion of other leading interictal biomarkers of the EZ – spikes, wideband HFOs, fast ripples, and ripples – at both the group and individual levels. To test these hypotheses, we analyzed an international multi-institutional dataset of intracranial recordings in subjects with known surgical outcome and at least 1 year of follow up using validated approaches to detect automatically each interictal biomarker.

## Methods

### Subject Selection

109 subjects with drug-refractory epilepsy who underwent intracranial phase II evaluation, surgical resection, and at least 1 year follow up were included from 4 tertiary epilepsy centers (site A: n = 39; site B: n = 12; site C: n = 44; site D: n = 14). We note that data from three prior studies evaluating HFOs as predictors of the epileptogenic zone were included in this study ^25-27^, in addition to any subsequently available datasets from subjects that met our inclusion criteria from these centers through 12/19/2021.

All enrolled subjects were scored using the International League Against Epilepsy (ILAE)^28^ scale to assess their surgical outcomes. For each subject, the resected volume (RV) was determined in direct consultation with the neurosurgeons, who identified which channels were resected by referring to the surgical records and comparing postoperative MRI with preoperative clinical labels. When evaluated in subjects who achieved successful seizure control after surgery (ILAE 1 subjects), the RV provides the best approximation of the EZ, though we note that even in these carefully selected subsets, the RV may contain more brain tissue than just the EZ (overlying tissue to reach deep lesions, for example).

All data were acquired with approval of the local IRBs and the Massachusetts General Hospital IRB.

### EEG Recording/Acquisition

All sites recorded with their local equipment. Data from site A and site B were recorded using a Harmonie acquisition system (Stellate, Montreal, Canada) with sampling rate of 2000 Hz. Data from site C were recorded using a Neuralynx acquisition system (Bozeman, MT, USA) with sampling rate of 32 kHz and a 9 kHz antialiasing filter. Data from site D were recorded using a Natus Quantum acquisition system with sampling rate of 4096 Hz and a 1.2 kHz anti-aliasing filter. Data from the Neuralynx and Natus Quantum acquisition systems were downsampled to 2500 Hz and to 2048 Hz, respectively, using the *decimate* function in MATLAB (The Mathworks, Natick, MA, USA). Chronic stereotactic-depth electrodes were used at all centers. Subdural grid and strip electrodes were used at sites A, C, and D. The site A and site B recordings included periods selected for slow wave sleep. The site C and site D recordings included approximately two hours of interictal data recorded from 1-3 AM.

We characterized intracranial electrode type as “depth” if stereotactically placed depth electrodes were used, or electrocorticography (“ECoG”) if subdural grid or strip electrodes were used. A subset of subjects with both depth electrodes and grid or strip electrodes were grouped based on which recording type accounted for most electrode contacts.

### EEG preprocessing

All data were referenced to bipolar differences between adjacent channels for a more localized representation of brain activity. An experienced electroencephalographer visually identified and manually removed channels with no signal or high noise. Periods of low-quality recordings caused by intermittent or continuous artifacts exceeding 30 seconds were also rejected, as well as all periods containing electrographic seizures.

To identify high amplitude artifacts, we detected all voltage peaks in the absolute value of the signal that exceeded 10 times the standard deviation of the mean peak amplitude for each channel and removed ± 1 second of data centered on each detected peak. To remove recording breaks, characterized by a discontinuous change in voltage, we computed the acceleration in voltage between adjacent data points for each 1 second non-overlapping epoch (an approximation of the second derivative) and epochs in which the difference between the maximum and minimum values exceeded 200 microvolts/s were removed.

### Automated ripple and fast ripple detection

For each subject from site A or B, the rates for interictal ripples and fast ripples were calculated using the methods described in ^25, 29^.

### Automated HFO detection

Wideband HFOs were detected on all data from all sites using the fully automated “quality HFO” (qHFO) algorithm ^30^ which detects wideband HFOs (80-500 Hz). This method has been validated to have similar yields as human reviewers. The EEG data were processed using MATLAB software (The Mathworks, Natick, MA, USA) and the General Data Flow Package ^30^.

### Automated spike detection

Spikes were detected on all data from all sites using Persyst 14 software (Persyst Development Corporation, San Diego, CA, USA) with the built-in Reveal algorithm ^31^, the most widely used commercially available spike detection software.

### Automated spike ripple detection

Our group has previously developed two separate spike ripple detectors for use in scalp EEG: a feature-based algorithm applied to time series data ^18, 32^, and a convolutional neural network (CNN) applied to spectrogram images ^33^. To apply these tools to intracranial data, we first retrained the CNN detector on hand-marked events. We then combined the two detectors to optimize precision suitable for prolonged, multichannel recordings and validated the resulting combined detector on hand-marked events using a leave-one-out cross-validation procedure. The details of these steps are outlined below.

To create a gold standard dataset of intracranial spike ripple events to train and validate the intracranial spike ripple detector, we compiled a diverse training dataset using a subset of data from 18 subjects across the 4 sites with ILAE 1 surgical outcomes. For each subject, one expert (WS and KW after consensus training) manually inspected 10 minutes of data from 5 EEG channels within the EZ and marked all spike ripple events (n = 1700, across all 18 subjects). We then applied the feature-based detector to 5 channels outside of the EZ for each subject to define 950 false positive detections across all subjects. Finally, we randomly selected 1600, 1 s samples from the 5 channels outside of the EZ across all subjects as true negative detections. Spectrogram images of 1 s epochs (either containing a true positive, false positive, or true negative detection) were generated to train the CNN detector ^33^. As in ^33^, we applied transfer learning to train the CNN (i.e., we retrained layers of an existing neural network architecture using the spectrogram image training data) ^34^. The CNN returns the probability of a 1 s epoch containing a spike ripple. To perform leave-one-out cross-validation, we trained the CNN on all but one subject, and then evaluated the performance of the combined detector, which used both the feature-based detector and the CNN detector, on the held-out subject at different CNN probability thresholds. We repeated this process for all subjects in the training dataset, and then aggregated the results to evaluate the overall performance of the combined detector. The threshold that yielded the best precision was selected to classify the data not used in training.

### Statistical Analysis

To test the hypothesis that the majority of detected spike ripples were removed in subjects seizure free after surgical resection, we utilized the previously reported metric, the event rate ratio ^25^. This ratio compares the sum of events detected in resected channels against those in unresected channels using the following equation:

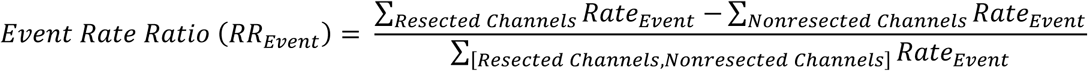

where the numerator is the difference between the summed event rates from removed channels and unremoved channels, the denominator is the summation of event rates from all channels, and all channels were observed for the same amount of time. The event rate ratio ranges from -1 to 1. Values greater than 0 in subjects with ILAE 1 outcome indicate that most events were removed and thus co-localized with the EZ, whereas values less than 0 indicate that the majority of events were not co-localized with the EZ (**Figure 1**). We tested if subjects seizure free after resection (ILAE 1 outcomes) had a higher proportion of events resected than non-resected (mean of the distribution of event rate ratio > 0, right tailed t-test), and repeated this test in the subsets of ILAE 1 subjects from the same site and with the same intracranial electrode type.

**Figure 1:**
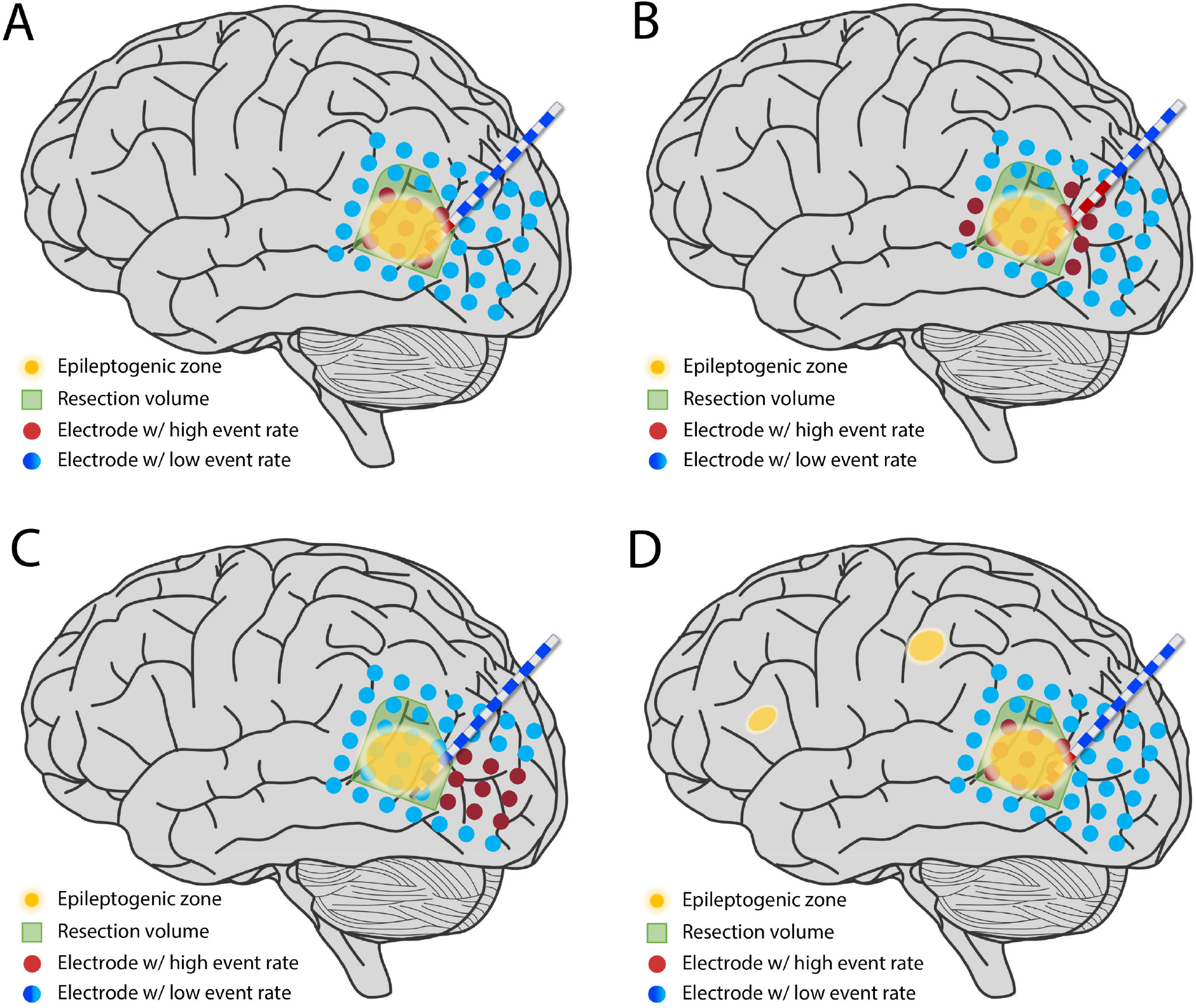
Illustrations of scenarios with different event rate ratios. ECoG (array of blue circles) and depth (white and blue line) electrodes monitor brain voltage. Channels with high event rates are indicated in red. The resected region (green area) overlaps with the true epileptogenic zone (yellow area) in subjects with good surgical outcomes. **(A)** Channels with high event rates are within the resected region, and the event rate ratio is 1. **(B)** Half of the channels with high event rates are within the resected region, and the event rate ratio is 0. **(C)** None of the channels with high event rates are within the resected region, and the event rate ratio is -1. **(D)** In some cases, the EZ may not be fully sampled by the intracranial investigation, resulting in a high event rate ratio but poor surgical outcome.

We next tested the hypothesis that subjects with surgical cure after resection had a higher proportion of spike ripples resected compared to subjects with continued seizures after resection (mean of the distribution of ILAE 1 event rate ratio > mean of the distribution of ILAE 2-5 event rate ratio, right-tailed t-test). To investigate further whether surgical outcome correlated with the spike ripple event rate ratio after accounting for potential confounding effects, we estimated a linear model of the spike ripple event rate ratio with five predictors: *Surgical Outcome =* {ILEA 1, ILEA 2-5}, the predictor of interest; *Recording Site* = {A,B,C,D}; *Recording Type* = {depth, ECoG}; *Duration* ={recording duration in minutes}; and a constant.

To test the hypothesis that a higher proportion of spike ripples were resected in subjects seizure free after surgery compared to other proposed biomarkers – spikes, wideband HFOs, fast ripples, and ripples – at the group level, we compared the event rate ratios of each biomarker (*RR*_*Spike Ripple*_, *RR*_*Spike*_, *RR*_*HFO*_, *RR*_*Fast Ripple*_, and *RR*_*Ripple*_) in subjects with ILAE 1 outcome by applying pairwise right-tailed exact binomial tests. We used this approach to account for cases with zero detections. To complement these group-level analyses, we also evaluated in subjects with ILAE 1 outcomes the proportion of channels with high event rates resected for each biomarker type. High event rates were defined as the 50th, 60th, 70th, 80th, 90th, and 95th quantiles of the event rate distribution for each biomarker.

To test this hypothesis at the individual level, we estimated surgical outcomes in individual subjects using each biomarker. Subjects with ILAE 1 outcomes were considered correctly predicted by the biomarker if the event rate ratio was positive (*RR*_*Event*_ > 0), indicating that the majority of events were removed. We tested if the success rate of estimated surgical outcomes was higher for spike ripples than other biomarkers (i.e., spike, wideband HFO, fast ripple, or ripple) using right-tailed two-proportion z-tests.

## Results

### The intracranial automated spike ripple combined detector has excellent precision

Given the high volume of multichannel data available in intracranial datasets, we developed a spike ripple detection strategy that optimizes precision. To do so, we required simultaneous spike ripple detection by both a feature-based detector that analyzes time series data and a convolutional neural network (CNN) detector that analyzes spectrogram images, which we previously developed ^32, 33^. Using a leave-one-out cross-validation procedure, we determined the probability threshold (0.65) of the CNN detector that achieved the highest precision of the combined detector against detections hand-marked by experts. While the combined detector identified less than half of hand-marked events (sensitivity, 42%), it had excellent precision (78.5%) and a low false positive rate (6.7%), resulting in balanced performance (F1 = 0.68). The combined detector required approximately 6.8 minutes to analyze 10 min from 10 channels of data sampled at > 2 kHz (1.8 min for the feature-based detector; 5 min for the CNN detector) on a single processor with 3.8 GHz CPU (64 GB RAM). Example intracranial spike ripples detected in the resected volumes of ILAE 1 subjects are shown in **Figure 2**.

**Figure 2:**
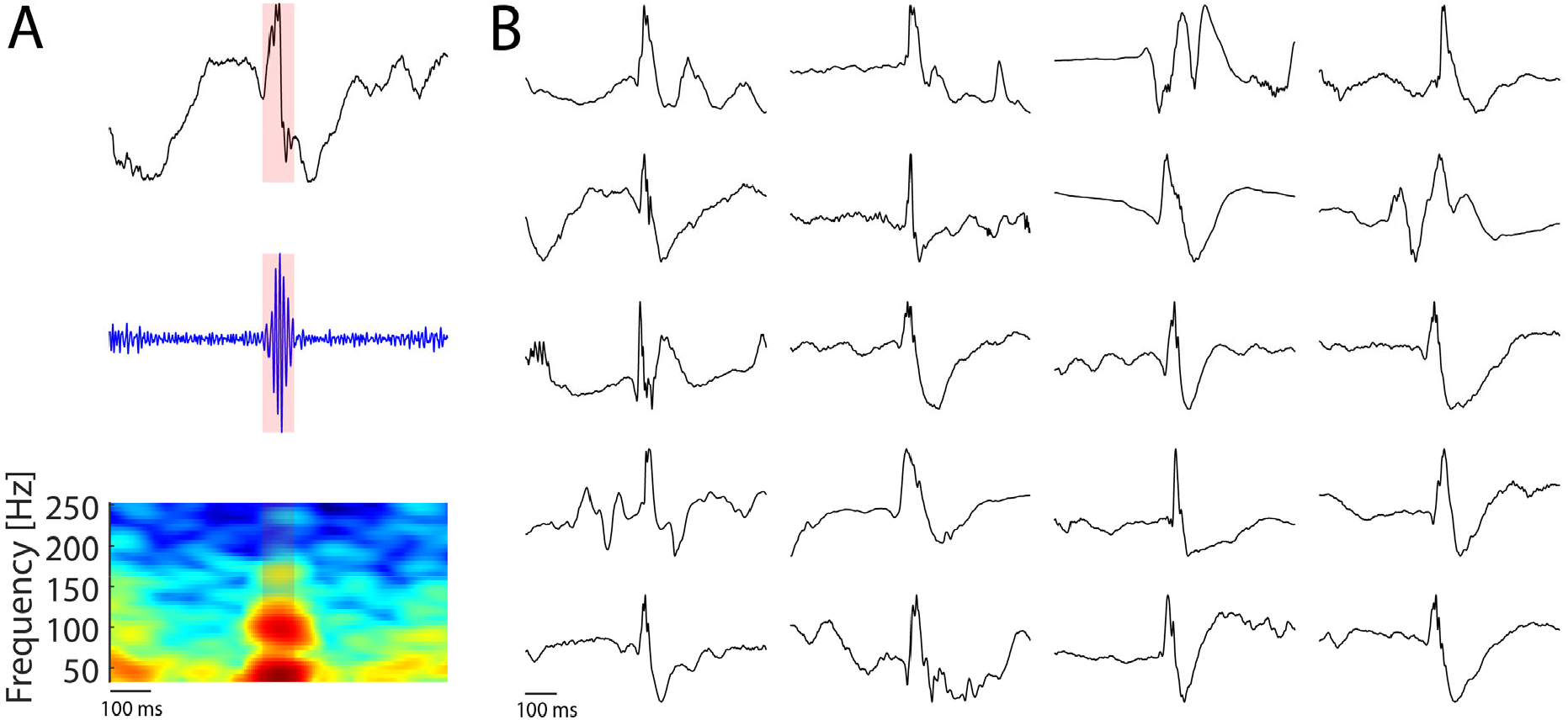
Example spike ripple detections. **(A)** Example detected spike ripple event, showing the unfiltered voltage recording (top), the 100-300 Hz bandpass-filtered voltage recording (middle), and the spectrogram (bottom). The spectrogram displays the power in decibels and warmer colors represent higher powers. Red shading in each subplot indicates the time interval of the detected spike ripple event. **(B)** Example unfiltered voltage recordings of spike ripple detections. All detections share the same feature of an epileptiform spike co-occurring with a ripple event.

### Data characteristics

Subjects with drug refractory epilepsy and known surgical outcomes after 1 year of follow up were included (n = 109, 59 females; median age 32 years, range 8-65 years). Among these, 48 subjects had ILAE 1 outcomes. Center-specific demographic information is provided in **Table 1**. The median duration of intracranial EEG data analyzed was 120 min/subject (range 10-121 min). Subject recordings included an average of 58 channels (range 11-147).

**Table 1.** Center-specific demographic information.

| Cohort | Number | Age mean (range, number of children) | Percent female |
| --- | --- | --- | --- |
| Site A | N = 39 | 31 years (8-62, 8) | 41% |
| Site B | N = 12 | 34 years (21-53, 0) | 92% |
| Site C | N = 44 | 34 years (11-65, 2) | 52% |
| Site D | N = 14 | 31 years (13-59, 5) | 64% |

B
|  | Class 1 | 2 | 3 | 4 | 5 |
| --- | --- | --- | --- | --- | --- |
| Site A | 19 | 3 | 7 | 5 | 5 |
| Site B | 3 | 0 | 2 | 3 | 4 |
| Site C | 16 | 4 | 7 | 4 | 13 |
| Site D | 10 | 4 | 0 | 0 | 0 |

### The majority of spike ripples were removed in subjects with surgical cure

The majority of spike ripple generating brain tissue was removed in drug-refractory epilepsy subjects who were seizure free after surgical resection (p = 1e-6, **Figure 3**). This result was qualitatively consistent at each site (Site A: p = 0.032; Site B: p = 0.092; Site C: p = 0.0005; Site D: p = 0.003) and in subjects with majority depth electrode recordings (p = 0.01) or majority subdural grid or strip recordings (p = 7e-6). Among ILAE 1 subjects, the mean spike ripple rate was higher in the RV (0.66/min, 95% CI [0.60, 0.72]) than in the non-removed tissue (0.06/min, 95% CI [0.07, 0.09], p = 3e-3).

**Figure 3:**
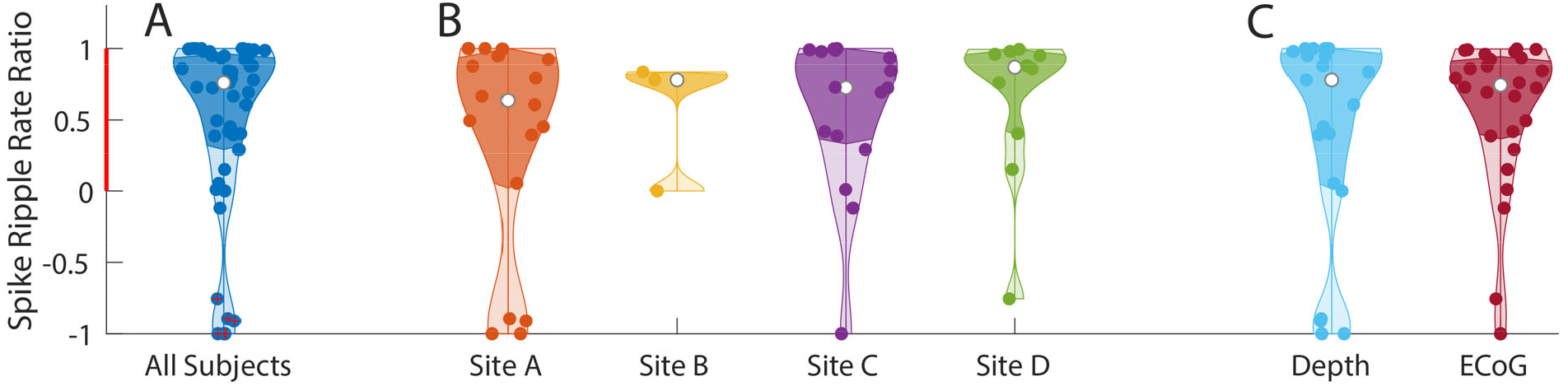
In subjects with good surgical outcome most spike ripples were removed. Distribution of the spike ripple rate ratio from **(A)** all subjects with ILAE 1 surgical outcome, **(B)** from each clinical site, and **(C)** from each intracranial electrode type. Each dot indicates one subject. Violin plots show the density of the spike ripple rate ratio values, the violin width represents the number of subjects at each value, the white dot represents the median value, and the darker shading indicates the interquartile range. Data points that deviate significantly from the majority of each data group are identified as outliers and marked with red crosses.

### More spike ripples were removed in subjects with surgical cure than in subjects with persistent seizures

Subjects with curative resection (ILAE 1) had a higher proportion of spike ripple generating brain tissue removed compared to those who were not seizure free (ILAE 2-5) (p = 0.02). This trend persisted after controlling for recording site, recording type, and duration (mean increase of the event rate ratio in ILAE 1 subjects compared to ILAE 2-5 subjects: 0.22, 95% CI [-0.04, 0.48], p = 0.099).

### Spike ripples identified the epileptogenic cortex better than other leading interictal biomarkers

Subjects with curative resection (ILAE 1) had a higher proportion of spike ripple-generating brain tissue removed compared to the proportion of removed tissue that generated spikes (p = 0.0004), wideband HFOs (p = 0.0004), fast ripples (p = 0.008), or ripples (p = 0.008, **Figure 4**).

**Figure 4:**
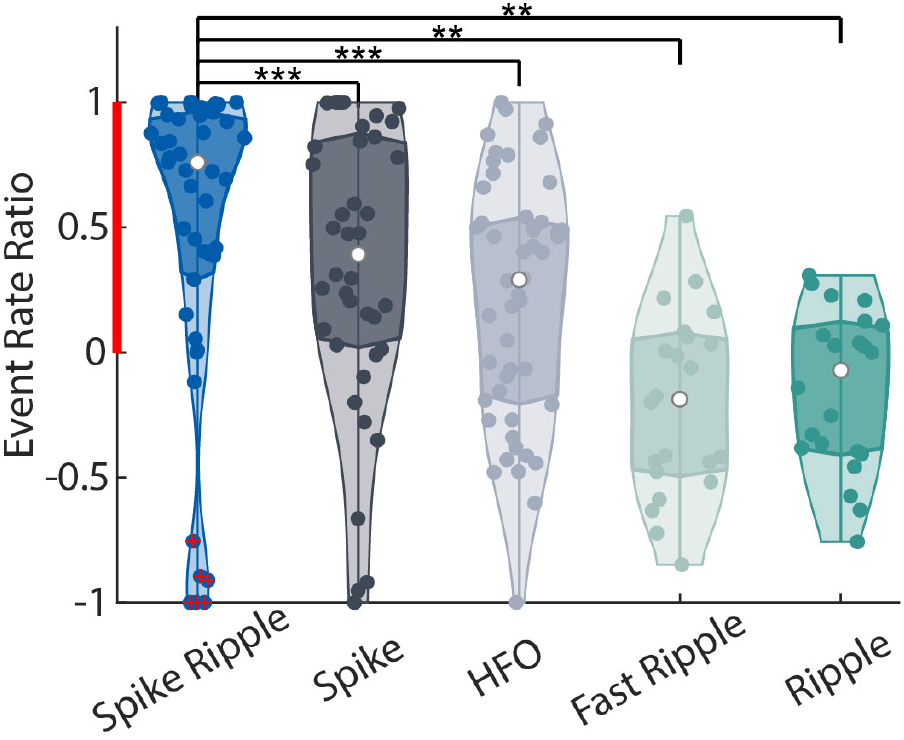
Spike ripples outperform other biomarkers in identifying the epileptogenic zone. Distributions of five different event rate ratios from subjects with ILAE 1 outcome (seizure free after surgery). Violin plots as in Figure 3. Among these subjects, spike ripples have a significantly higher event rate ratio than spikes, wideband HFOs, fast ripples, or ripples. Significance differences indicated as: ** p < 0.01; *** p < 0.001.

Although the RV in subjects with surgical cure after resection provides the best available estimate of the EZ, the RV may also include channels not necessary for surgical cure. Thus, to complement our analysis, we also evaluated whether the majority of channels with high event rates were resected. To do so, we calculated the proportion of channels with the highest event rates resected at thresholds (50^th^ to 95^th^ quantiles) for each subject. We found that the mean proportion of channels with the highest spike ripple rates resected exceeded the mean proportion of channels with the highest spike, wideband HFO, fast ripple, or ripple rates resected, in all quantiles evaluated (**Table 2**). Together, these findings provide strong support that spike ripple is a more specific biomarker for the epileptogenic zone than other leading interictal biomarkers.

**Table 2.**
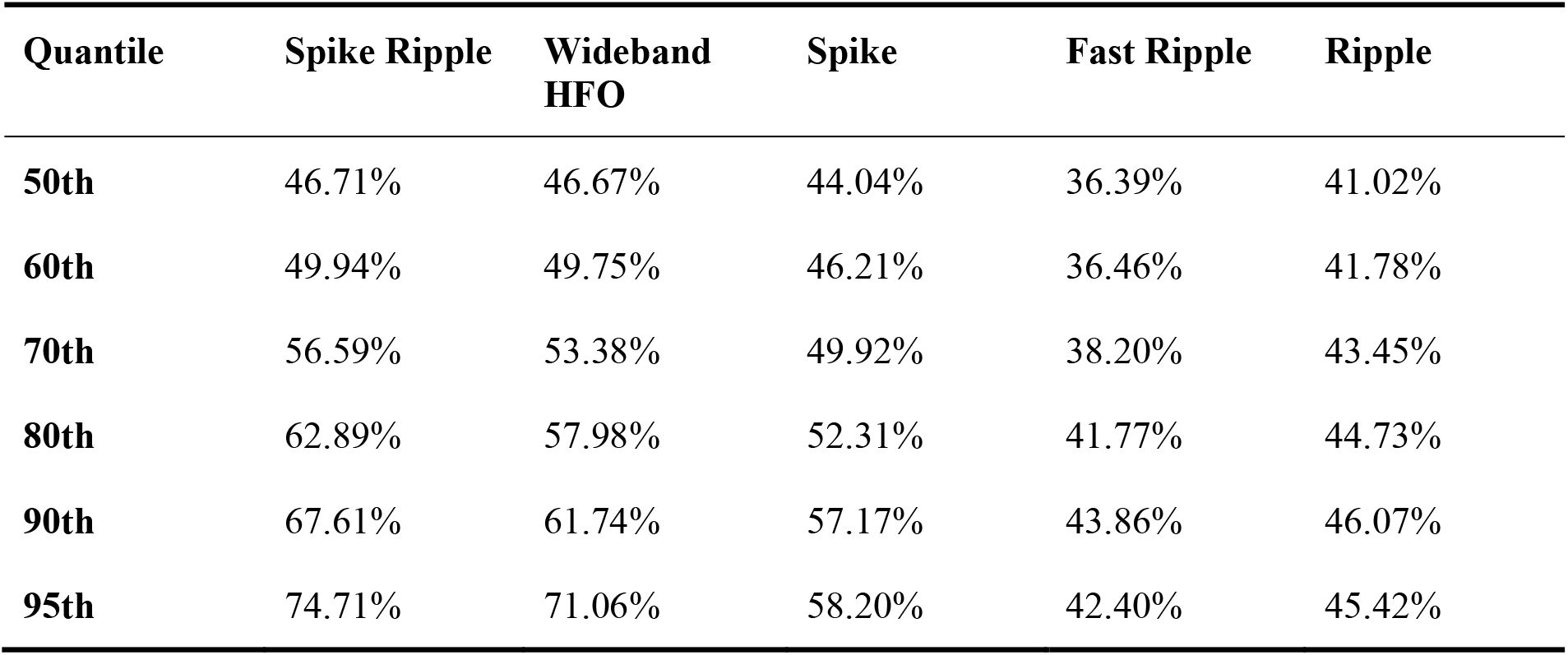
Mean proportion of resected high-rate channels for each interictal biomarker type in ILAE 1 outcome subjects at different thresholds.

| Quantile | Spike Ripple | Wideband HFO | Spike | Fast Ripple | Ripple |
| --- | --- | --- | --- | --- | --- |
| **50th** | 46.71% | 46.67% | 44.04% | 36.39% | 41.02% |
| **60th** | 49.94% | 49.75% | 46.21% | 36.46% | 41.78% |
| **70th** | 56.59% | 53.38% | 49.92% | 38.20% | 43.45% |
| **80th** | 62.89% | 57.98% | 52.31% | 41.77% | 44.73% |
| **90th** | 67.61% | 61.74% | 57.17% | 43.86% | 46.07% |
| **95th** | 74.71% | 71.06% | 58.20% | 42.40% | 45.42% |

### The majority of spike ripples were removed in most subjects with seizure freedom after surgery

If a biomarker identifies the EZ, we expect its removal to correlate with successful resective surgery. Consistent with this expectation, at the individual subject level, we found that the majority of spike ripples were removed (i.e., *RR*_*Spike Ripple*_ > 0) in 83% (40 of 48) of subjects with ILAE 1 outcome. The proportion of ILAE 1 subjects with the majority of spike ripples removed was higher than the proportion of subjects with the majority of spikes (69%, p = 0.04), wideband HFOs (63%, p = 0.009), fast ripples (36%, p = 2e-5), or ripples (45%, p = 0.0007) removed.

Although the majority of spike ripples were removed in 40 of 48 subjects with ILAE 1 outcome, the biomarker failed in eight subjects. To evaluate possible causes of biomarker failure, we further analyzed the spike ripple results for these eight subjects. In four subjects, the spike ripple rates were low, with less than 10 spike ripples detected across all channels in each case. In these four subjects, the median spike ripple rate was 0.075/min (mean 0.125/min, range 0-0.27/min). In contrast, the median spike ripple rate in the RV of the other 44 subjects was 1.7/min (mean 4.4/min, range 0.033-28.2). In three subjects, relatively high rates of spike ripples (> 1.1/min) were detected in a hippocampus that was not resected. In two of these cases, the hippocampi were clinically determined to be part of the seizure onset zone, but ultimately resections sparing the hippocampi were pursued and the patients achieved 1 year seizure freedom. In one subject, we found no apparent explanation for biomarker failure.

Overall, these results suggest that spike ripples provide a better estimate of the EZ than other leading interictal biomarkers. However, if spike ripples are detected with low rates across all channels, or at high rates in hippocampi, the biomarker may be less accurate.

## Discussion

We evaluated whether spike ripples, the combination of epileptiform spikes and ripples, accurately localized to the epileptogenic zone in an international cohort of subjects who underwent intracranial investigation, surgery, and post-surgical follow up. We found that the majority of spike ripples were removed in subjects who became seizure free after surgical resection, and that this finding was consistent across different epilepsy centers and recording electrode types. Additionally, we found that a higher proportion of spike ripples were removed in subjects who were seizure free after resection than in those who were not. Finally, we found that a greater proportion of spike ripples were removed in subjects that were seizure free after resection than the proportion of spikes, wideband HFOs, fast ripples, and ripples. Overall, we conclude that spike ripples have better specificity for epileptogenic tissue than other leading interictal biomarkers and therefore provide a valuable tool for identifying the epileptogenic zone to guide surgical resection.

Previously, we have shown good intra-rater reliability and performance using semi-automated ^32^ and fully-automated ^18, 33^ techniques on scalp EEG data. Here we introduce a combined detector that extracts both time series features and spectral information to detect spike ripples in intracranial data. We find that this detector has high precision in identifying spike ripples on intracranial EEG recordings. In prolonged, multichannel intracranial recordings, this detector has sufficient sensitivity to detect nearly half of all events and excellent precision to limit false detections. This novel automated approach enables objective quantification of spike ripples, providing a reproducible approach to estimate the EZ.

To assess biomarker performance, we measured the event rate ratio, the same metric applied in previous studies to evaluate biomarkers for the EZ, including a prospective clinical trial ^25, 36, 37^. We note that large resection volumes can falsely inflate this metric, but this concern is mitigated by the inclusion of multiple centers and surgical approaches and the comparison of different biomarkers using the same metric. Although spike ripples provided a reliable biomarker for the EZ in most subjects studied here, the majority of automatically detected events occurred outside the EZ in eight subjects. Among these subjects, four were found to have low event detection rates among ILAE 1 subjects. This finding suggests that accurate detection of the EZ using this biomarker may require a minimum number of detected events. Based on the data available here, we suggest that recording durations should be sufficient to obtain > 10 detections, where higher rates are more reliable. Although the majority of hippocampal EZ were identified correctly using the spike ripple rate ratio (9 of 9 subjects with ILAE 1 outcome and hippocampal resection), the biomarker failed in three other subjects with ILAE 1 outcome who had high spike ripple detection rates in hippocampi that were not resected. These results may indicate that the unresected hippocampus may in fact be epileptogenic if the subject were followed for a longer period of time ^38^ or alternatively that hippocampal sharp wave ripples, which share many features with spike ripples, were misidentified as spike ripples by the detector ^35, 39^. Until validated methods to distinguish spike ripples from sharp wave ripples are available, limiting spike ripple evaluations to non-hippocampal tissue may be the most prudent. Excluding subjects with low spike ripple rates (< 10 total or < 0.3 detections per minute) and those with high spike ripple rates in hippocampus only, spike ripples accurately localized the EZ in 40 of the remaining 41 subjects, indicating that spike ripples are a highly reliable cortical biomarker when present.

We found a stronger relationship between spike ripples and the EZ compared to other biomarkers (spikes, wideband HFOs, fast ripples, or ripples) and the EZ. Whether spike ripples reflect a unique event compared to spikes or ripples alone is not yet understood. The observation that spike ripples better localize to the EZ than spikes suggests that these events may be originating rather than propagated discharges. In humans, spike ripples correlate with higher single-unit firing rates compared to spikes ^40^. However, the roles of different cell types (e.g., interneurons) in spike ripple generation remain unclear ^41, 42^. Understanding the cellular mechanisms that support spike ripples, and whether treating these mechanisms impacts seizure recurrence, remains an open challenge.

When interpreting the results reported here, we note the following limitations. First, the epileptogenic zone is a theoretical concept and may not exactly match the resected volume in subjects with good surgical outcomes, as utilized here. To mitigate this concern, we used the best available measure of the EZ, the resected volume in patients with surgical cure. Second, we analyzed data from 1-3 AM as a proxy for non-rapid-eye-movement (NREM) sleep for 2 of 4 sites, as sleep scoring was not available. Although previous analysis indicated little difference between interictal 1-3 AM data and interictal NREM sleep data, definitive NREM recordings may be preferable ^8, 48^. Finally, we utilized data collected prospectively in a multicenter study evaluating HFOs to perform a retrospective analysis. Future work evaluating the impact of spike ripples on surgical outcome in a prospective cohort is required to validate these findings.

In conclusion, this study provides evidence that spike ripples are an improved biomarker for detecting epileptogenic tissue compared to spikes, wideband HFOs, fast ripples, and ripples.

## Data Availability

Data produced in the present study are available upon reasonable request to the relevant co-authors from each institution.

## Acknowledgments

This work was supported by NIH NINDS R01NS119483. The authors would like to acknowledge Daniel Lachner Piza, Rina Zelmann, and Vasileios Kokkino for supporting data acquisition; Uhrich Summer for assistance with spike detection; and Jean Gotman for discussion of the statistical method and the future directions.

## Author Contributions

MAK and CJC contributed to the conception and design of the study. SVG, JJ, BHB, GAW, and WCS contributed to data acquisition. WS, CJC, MAK, KGW, DS, XH, UTE, and RMR contributed to data analysis. WS wrote the first draft of the manuscript. All authors edited and approved the manuscript.

